# Effectiveness of 2^nd^ Skull Products in Reducing Head Impact in Simulated Sports

**DOI:** 10.1101/2020.05.05.20092569

**Authors:** Marina Ritchie, Honghuang Lin, Rhoda Au

**Affiliations:** Department of Anatomy & Neurobiology, Boston University School of Medicine, Boston, MA 02118, USA; Department of Neurology, Boston University School of Medicine, Boston, MA 02118, USA; Framingham Heart Study, National Heart, Lung, and Blood Institute, Boston, MA 01702, USA; Department of Epidemiology, Boston University School of Public Health, Boston, MA 02118, USA; Section of Computational Biomedicine, Department of Medicine, Boston University School of Medicine, Boston, MA 02118, USA

**Keywords:** head impact, traumatic brain injuries, contact sports, protective headgears, concussion

## Abstract

Cumulative effects of repetitive head impact have been highly associated with short and long term neurological conditions. Despite high rates of head injuries, many people in the U.S. and around the world continue to participate in collision sports. As one approach to address this concern, 2^nd^ Skull has developed supplemental protective headgears including a skull cap, scrum cap and headband that can be utilized in both helmet and non-helmet sports. To test the effectiveness of 2^nd^ Skull products as an adjunctive tool to reduce the severity of head impact, data were extracted from a series of tests completed at four sites under laboratory conditions using linear, projectile and rotational impactor to simulate blows to the head in sports. The majority of test cases showed the same pattern of reduced impact severity with the addition of 2^nd^ Skull padding. It remains to be seen if these laboratory results will translate to the field.

## Introduction

In the past five years, there has been a surge of interest in literature and media surrounding sports-related injuries and their long-term consequences. More specifically, contact sports have been a topic of considerable discussion due to the pace of neuropathological studies evidencing chronic traumatic encephalopathy (CTE) in subsets of populations with high exposure to collision and impact (Bieniek et al., 2020; Mckee et al., 2009; Mez et al., 2020). While popularity and participation have not deflated, the increased risk of traumatic brain injuries (TBI) associated with sports raises the issue of whether this is a public health concern. The most common form of TBI as a recognized medical condition is a concussion, which is currently defined by evidence of a blow to the head that results in symptoms most often lasting from a few hours to a few weeks but can persist for months or even years (Shaw et al., 2002). Consequences of concussions are of great concern in sports-related contexts, with 1.6 to 3.8 million athletes in the US experiencing sports-related concussions each year, contributing to large healthcare expenditures (Langlois et al., 2006; Marshall, 2012). There is wide variability in acute symptoms of mild TBI (mTBI) ranging from headaches, dizziness, blurred or doubled vision, and memory loss (Prince & Bruhns, 2017). While most symptoms dissipate quickly, they may develop into chronic conditions and cause neurodegeneration (Cruz-Haces et al., 2017; Mez et al., 2017). Longitudinal studies have suggested that history of TBI can have pathological long-term effects such as an increased risk of Alzheimer’s disease (AD) and CTE (Fleminger et al., 2003; Gavett et al., 2011). Even without the loss of consciousness, repetitive head impacts (RHI) and sub-concussive blows have acute and chronic consequences including structural and functional changes to the brain (Mainwaring et al., 2018; Davenport et al., 2016).

Conditions due to repetitive head trauma were first noticed in former boxers in the1920s and was commonly referred to as “punch-drunk syndrome” (Martland, 1928). Now punch-drunk syndrome is commonly associated with CTE or other TBIs. CTE is a neurodegenerative disease associated with exposure to repetitive head impacts (RHI), like those incurred during American football. In a study conducted by Mez and colleagues, the brains of 202 deceased American football players were studied for neuropathological evidence of CTE (Mez et al., 2017). Evidence of CTE was found in 87% of the brains, suggesting that long-term neurological conditions may be related to sports-related head impact. Furthermore, 47% of the studied brains indicated severe neurodegenerative pathology that resulted in dementia-related or Parkinsonian-related death amongst these individuals (Mez et al., 2017). Along with AD and CTE, these injuries can result in behavioral changes, post-concussive syndrome, and motor neuron diseases which are disproportionate to typical age-related changes (Babcock et al., 2013; Cole et al., 2015; Robbins et al., 2014). Despite the accumulating recognition of associations between repetitive head injuries and neurological disorders, little is known about effective treatment approaches, which highlight the importance of risk reduction and potential prevention (Mez et al., 2017; Maas et al., 2017).

Preventative headgears made to reduce head injury risk have been tested and implemented on a large scale in professional and semi-professional sports including the National Football League. Special attention has been paid to helmets, with numerous prototypes undergoing rigorous testing to determine models that optimally reduce head impact severity (Bartsch et al., 2012; Bottlang et al., 2020; Viano et al., 2006). While other types of protective headgear have also been evaluated, the scientific consistency and validity have predominantly been reported from those that are helmet based (Sone et al., 2017). Cutting edge helmets, however, are professional or semi-elite focused with limited data on adjunctive products and are not readily available at the general population level.

To address this issue, 2^nd^ Skull Inc. has developed novel caps which incorporate a thin layer of impact management material into a traditional under-helmet liner cap. While many athletes choose to wear thin liner caps underneath these helmets for reasons such as sun protection and sweat management, 2^nd^ Skull’s scientifically engineered 2mm Skull Cap with linear, projectile and rotational protection can add a layer of supplementary insulation to the helmets without changing the existing safety protocol. Furthermore, 2^nd^ Skull has also developed headbands and scrum caps for non-helmet sports making it scalable to a broader population. To investigate the effectiveness of 2^nd^ Skull products as an adjunctive buffer against impact, test series at independent third-party laboratories were conducted. Results obtained from the trials were consolidated and quantitatively reviewed to further evaluate the efficacy and overall pattern across all sites.

The objective of this study is to examine the effect of 2^nd^ Skull products on the protection against impacts. The results from multiple independent laboratories were evaluated and compared.

## Method

### 2^nd^ Skull products

In contact sports, the addition of helmets has generally been associated with larger reduction in response metrics of impact that reach the head. However, protective headgear often pose a series of trade-offs between safety, comfort and other sport-specific parameters (Graham et al., 2014). The adaptable characteristics of 2^nd^ Skull products, however, are engineered to improve energy management of impact for use with a variety of helmet designs (i.e. single hit versus multiple-impact helmets) as well as absorb impact for non-helmet sports. Both the cap and headband have a thin layer of lightweight poron XRD made from urethane molecules that are soft and flexible at rest but harden under sudden pressure (2^nd^ Skull, n.d). This mechanism has been developed for protection against linear impact from direct hits. Whereas previous reports have focused on linear impact in contact sports, recent studies experimentally investigating the effect of rotational impact have proposed rotational kinematics to be a better indicator of TBI risk (Kleiven, 2013). In response to current data, 2^nd^ Skull products have added an independent layer between the head and helmet, which creates a slip pane upon rotation impact. It has been hypothesized that this structure will reduce the amount of energy transferred to the head. The lightness of the cap also protects the head from rotational acceleration. While thicker and heavier layers can protect the head from linear acceleration, added mass and radius to the head can result in increased rotational acceleration (Graham et al., 2014).

### Experimental tests

To test the efficacy of risk reduction, independent third-party laboratories were commissioned by 2^nd^ Skull to complete a series of tests that adhered to the National Operating Committee on Standards for Athletic Equipment (NOCSAE) and American Society for Testing and Materials (ASTM) standards for new helmets. Three products, 2mm 2^nd^ Skull cap, 4mm headband, and rugby scrum cap were evaluated under laboratory conditions using linear, projectile and rotational impactor to simulate head hits in sports. For helmet-based sports and activities (American football, hockey, lacrosse, softball, baseball, ski & snowboard, BMX, cycling, skateboard and equestrian), helmet testing featured standard unprotected head forms and head forms with the addition of 2^nd^ Skull caps to measure variances in severity index, acceleration and peak (g) improvement. For non-helmet based sports (soccer and rugby), standard head forms were tested against head forms with 2^nd^ Skull headgear (headband and scrum cap) to measure variances in force improvement. There are subtle differences in testing protocol depending on the laboratory and model including impact velocity and location of impact. For tests with more than one trial, the average results of the same test protocol were reported.

Tests were completed in the following four laboratories:

- Chesapeake Testing (a division of NTS) assessed 2^nd^ Skull caps with baseball, football, hockey, lacrosse and softball helmets. Testing was achieved through use of a Modular Elastic Programmer (M.E.P.) helmet pad and Hybrid III head and neck on a linear impactor.
- ICS Inc. Laboratories completed a custom research and development testing per 2^nd^ Skull’s specifications simulating impact in hockey, ski/snowboard, lacrosse and soccer. 2^nd^ Skull products were stored for a minimum period of 4 hours at a constant temperature of 72°F ±5°F (22°C ± 2°C),
- Dynamic Research, Inc (DRI) conducted tests using 2^nd^ Skull caps for linear and rotational acceleration in simulated equestrian and rotational kinematics in simulated hockey.
- Biokinetics conducted additional football simulation tests with three different types of football helmets including Vicis Zero1 (VZ1), Riddell Speed Classic (RSC), and Schutt Vengeance Pro (SVP). The testing was conducted using Biokinetics’ linear impactor and followed the NFL linear impactor helmet test protocol.
  - Averaged results of different helmet location and drop velocities of 5.5m/s, 7.4m/s and 9.3m/s were reported.

Please refer to the **Supplemental Material** for more information on each test.

### Statistical Analysis

To obtain the percent improvement (%), the scores from helmet with 2^nd^ Skull product were divided by the helmet only measures.

## Results

The test series were conducted across four independent laboratories that simulated linear, projectile and rotational impact from sports. The list of simulated sports, test sites and impact severity can be found in **Table 1**. The additional 2^nd^ Skull padding showed a reduction of impact in the majority of simulations, however in some cases, no improvement or worse outcomes were also recorded. Percent improvement ranged from −20% to 60%. Mean scores were used to calculate the percent of improvement for each area of impact as the number of test trials for each simulation type varied between 1 and 4. A summary of results for all tests are included in **Table 2**.

**Table 1:**
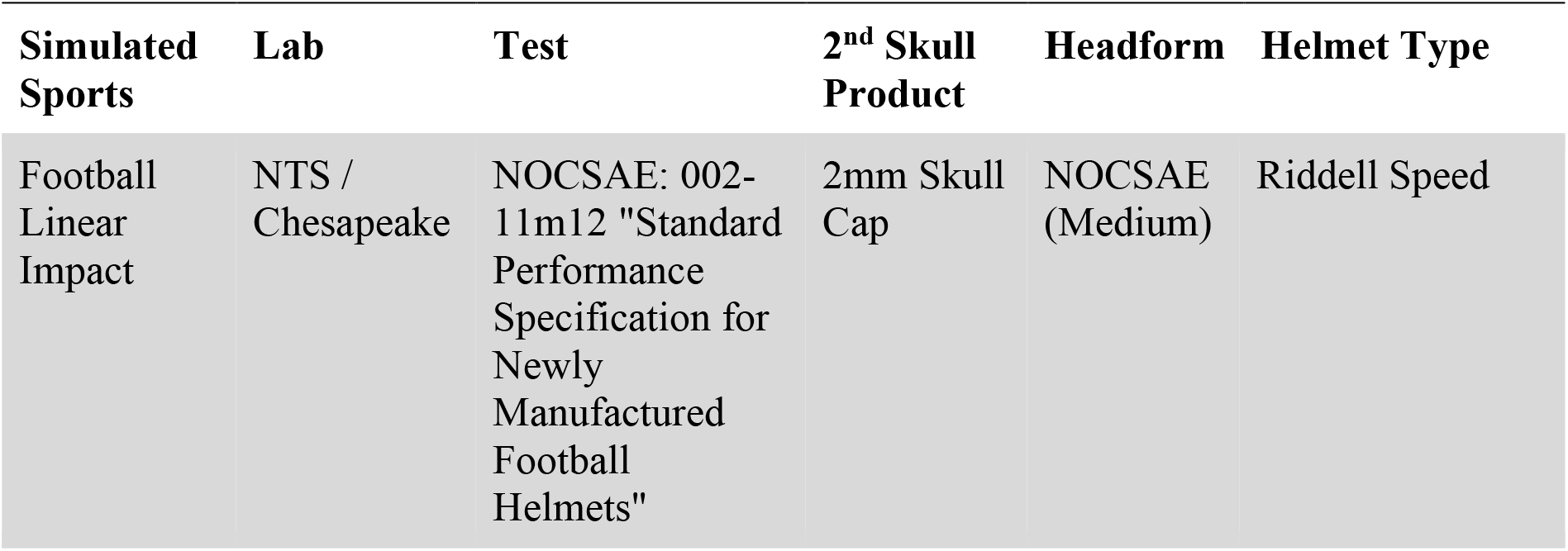
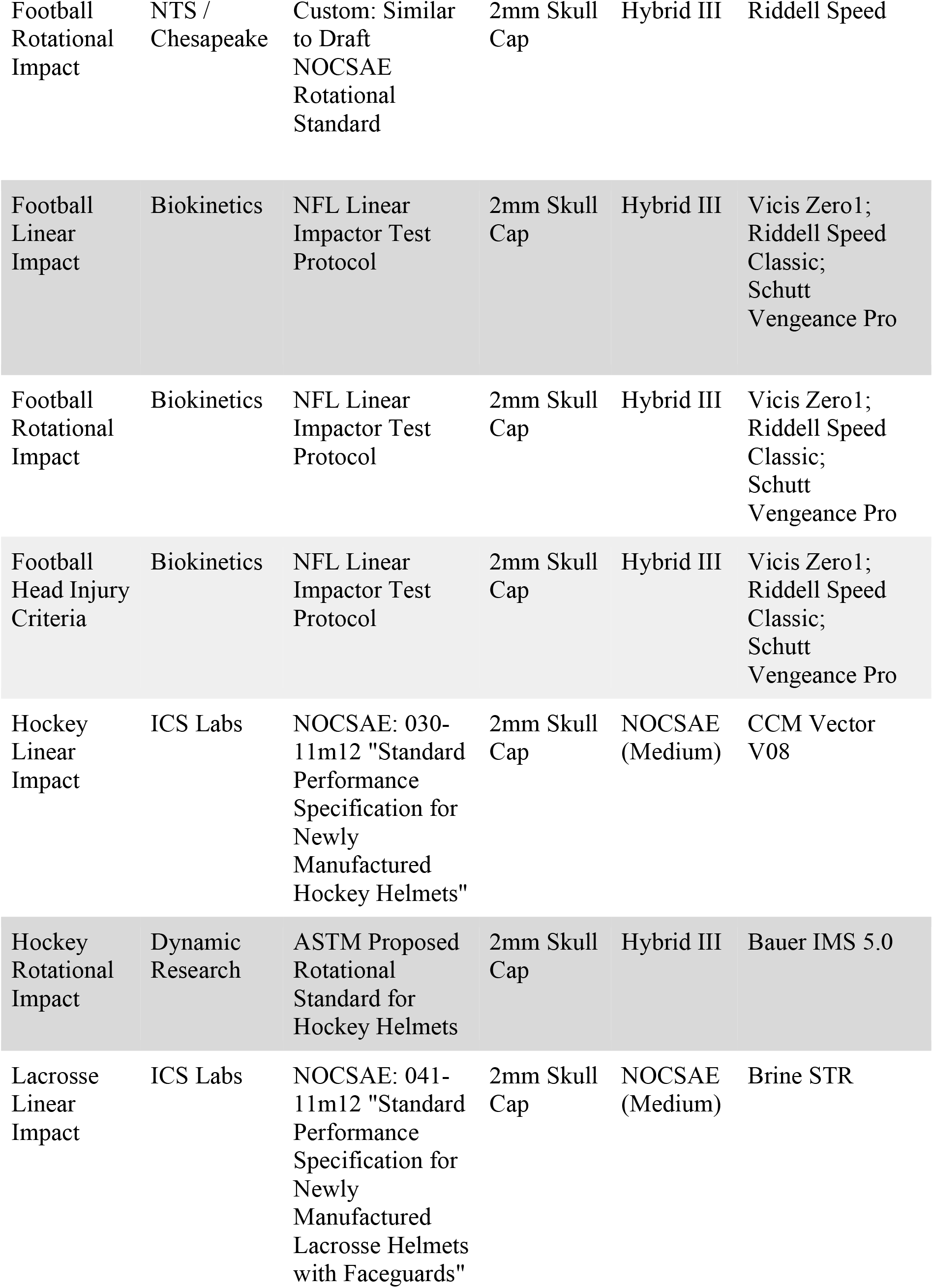
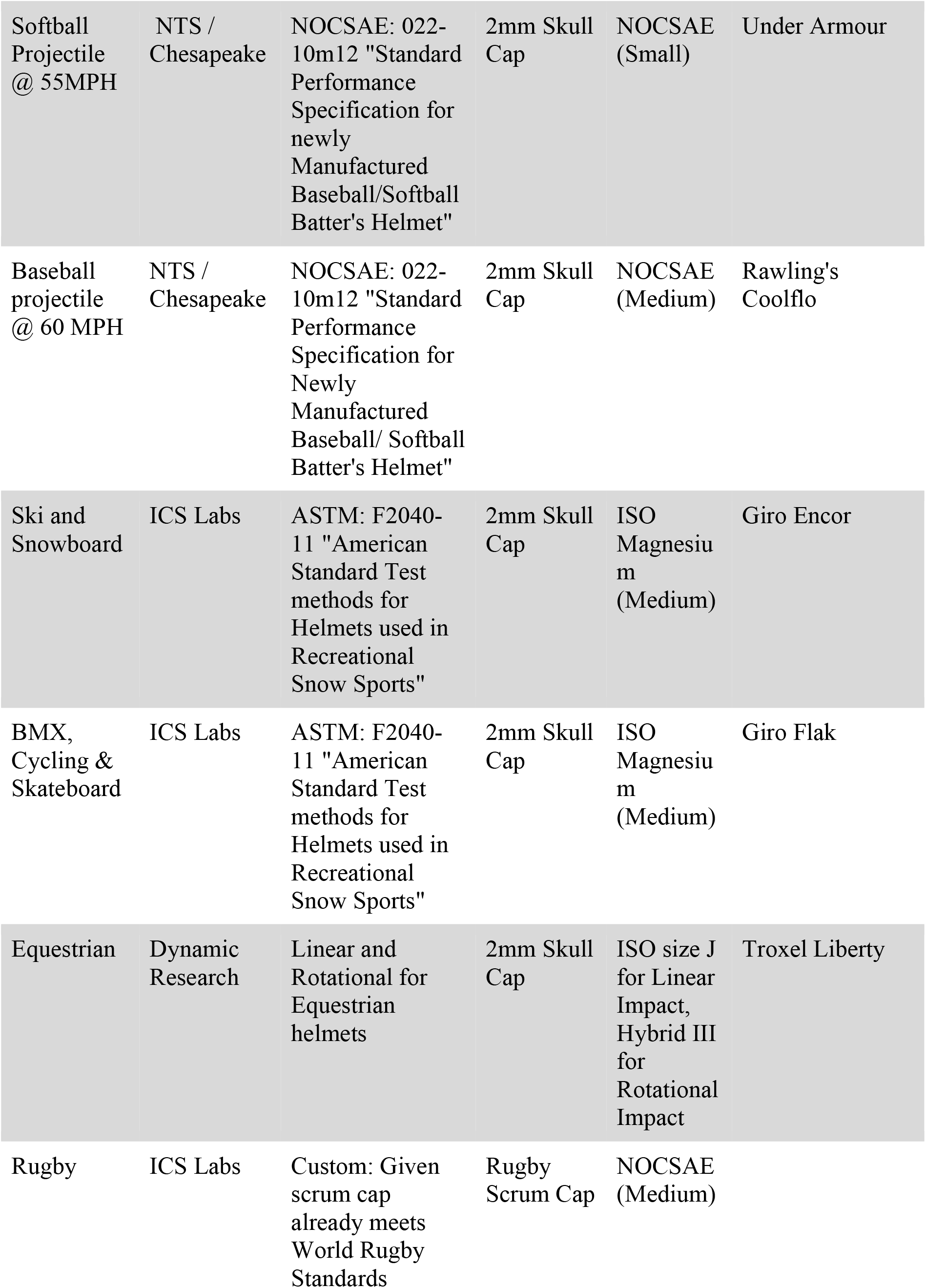
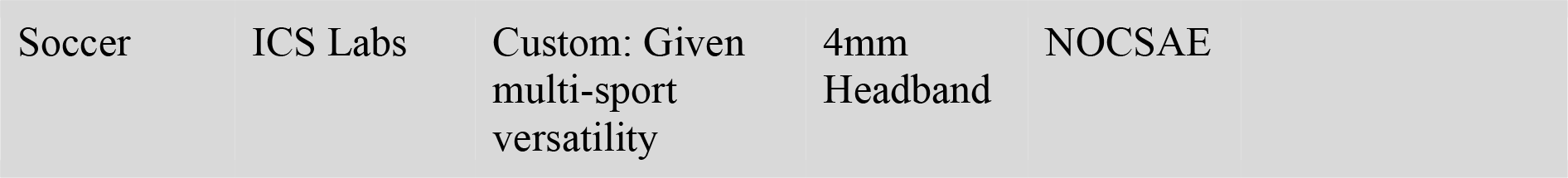
List of simulated sports and tests used to assess impact severity.

| Simulated Sports | Lab | Test | 2<sup>nd</sup> Skull Product | Headform | Helmet Type |
| --- | --- | --- | --- | --- | --- |
| Football Linear Impact | NTS / Chesapeake | NOCSAE: 002-11m12 "Standard Performance Specification for Newly Manufactured Football Helmets" | 2mm Skull Cap | NOCSAE (Medium) | Riddell Speed |
| Football Rotational Impact | NTS / Chesapeake | Custom: Similar to Draft NOCSAE Rotational Standard | 2mm Skull Cap | Hybrid III | Riddell Speed |
| Football Linear Impact | Biokinetics | NFL Linear Impactor Test Protocol | 2mm Skull Cap | Hybrid III | Vicis Zero1; Riddell Speed Classic; Schutt Vengeance Pro |
| Football Rotational Impact | Biokinetics | NFL Linear Impactor Test Protocol | 2mm Skull Cap | Hybrid III | Vicis Zero1; Riddell Speed Classic; Schutt Vengeance Pro |
| Football Head Injury Criteria | Biokinetics | NFL Linear Impactor Test Protocol | 2mm Skull Cap | Hybrid III | Vicis Zero1; Riddell Speed Classic; Schutt Vengeance Pro |
| Hockey Linear Impact | ICS Labs | NOCSAE: 030-11m12 "Standard Performance Specification for Newly Manufactured Hockey Helmets" | 2mm Skull Cap | NOCSAE (Medium) | CCM Vector V08 |
| Hockey Rotational Impact | Dynamic Research | ASTM Proposed Rotational Standard for Hockey Helmets | 2mm Skull Cap | Hybrid III | Bauer IMS 5.0 |
| Lacrosse Linear Impact | ICS Labs | NOCSAE: 041-11m12 "Standard Performance Specification for Newly Manufactured Lacrosse Helmets with Faceguards" | 2mm Skull Cap | NOCSAE (Medium) | Brine STR |
| Softball Projectile @ 55MPH | NTS / Chesapeake | NOCSAE: 022-10m12 "Standard Performance Specification for newly Manufactured Baseball/Softball Batter's Helmet" | 2mm Skull Cap | NOCSAE (Small) | Under Armour |
| Baseball projectile @ 60 MPH | NTS / Chesapeake | NOCSAE: 022-10m12 "Standard Performance Specification for Newly Manufactured Baseball/ Softball Batter's Helmet" | 2mm Skull Cap | NOCSAE (Medium) | Rawling's Coolflo |
| Ski and Snowboard | ICS Labs | ASTM: F2040-11 "American Standard Test methods for Helmets used in Recreational Snow Sports" | 2mm Skull Cap | ISO Magnesium (Medium) | Giro Encor |
| BMX, Cycling & Skateboard | ICS Labs | ASTM: F2040-11 "American Standard Test methods for Helmets used in Recreational Snow Sports" | 2mm Skull Cap | ISO Magnesium (Medium) | Giro Flak |
| Equestrian | Dynamic Research | Linear and Rotational for Equestrian helmets | 2mm Skull Cap | ISO size J for Linear Impact, Hybrid III for Rotational Impact | Troxel Liberty |
| Rugby | ICS Labs | Custom: Given scrum cap already meets World Rugby Standards | Rugby Scrum Cap | NOCSAE (Medium) |  |

|  |  |  |  |  |
| --- | --- | --- | --- | --- |
| Soccer | ICS Labs | Custom: Given multi-sport versatility | 4mm Headband | NOCSAE |

**Table 2:**
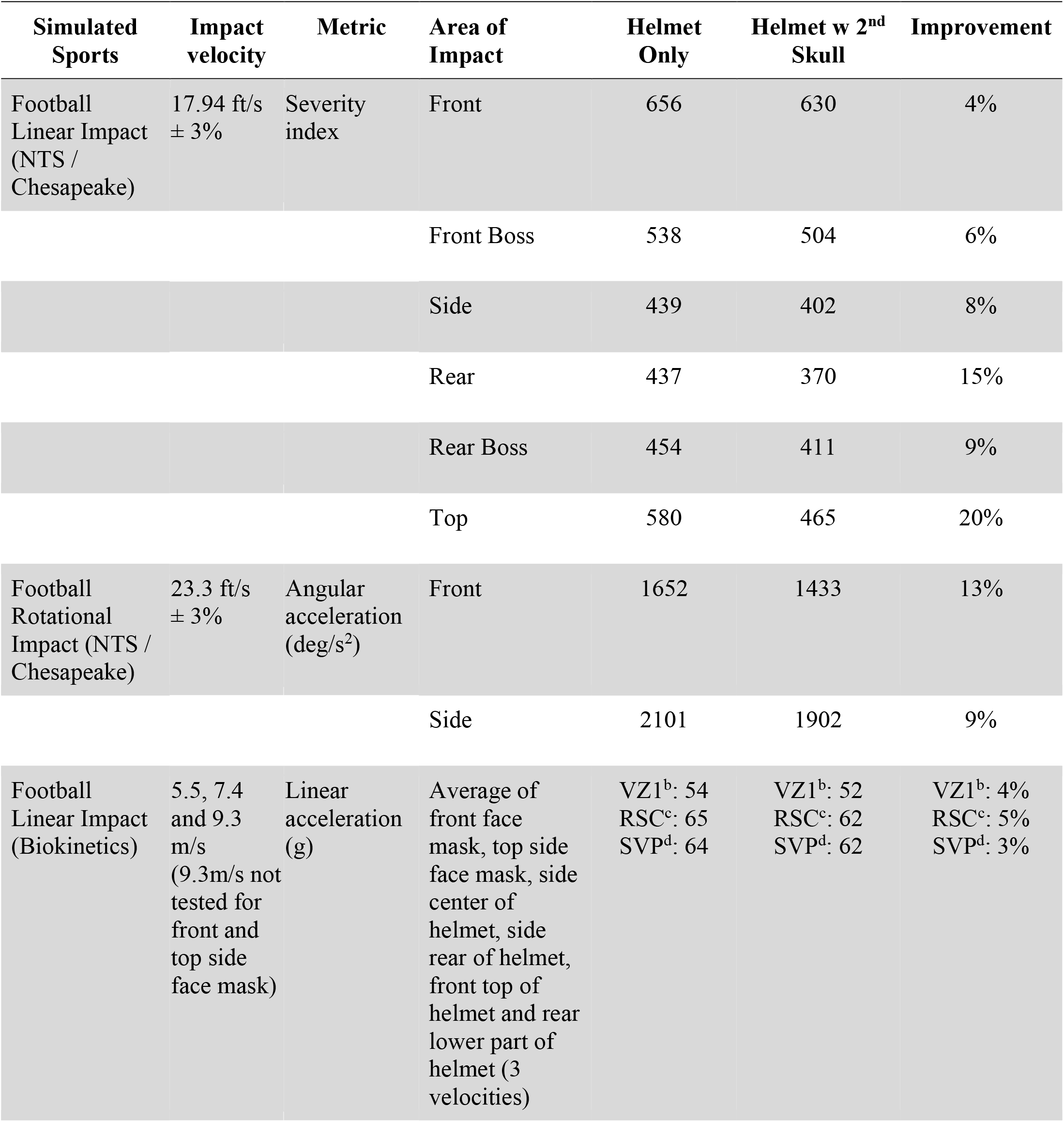
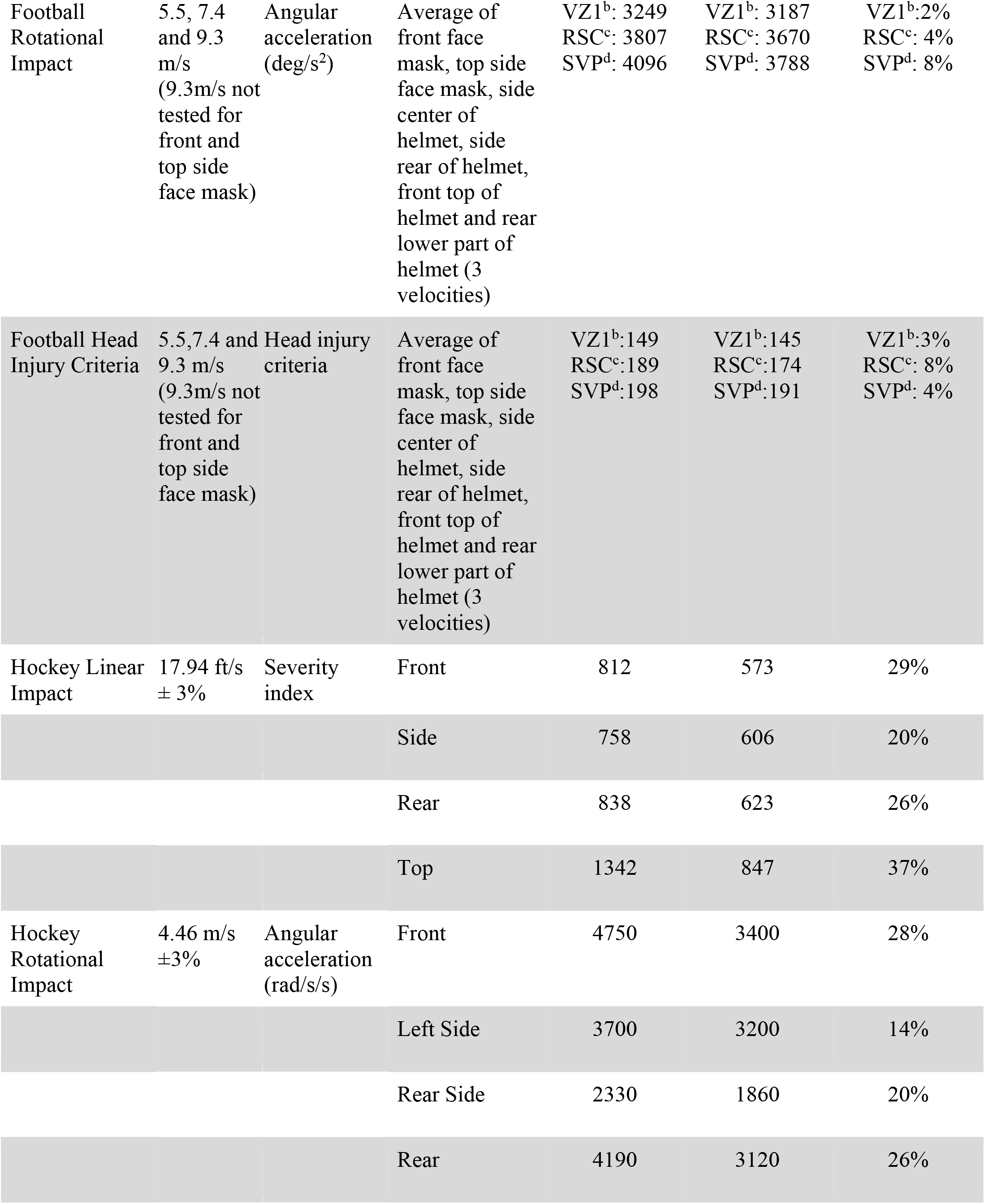
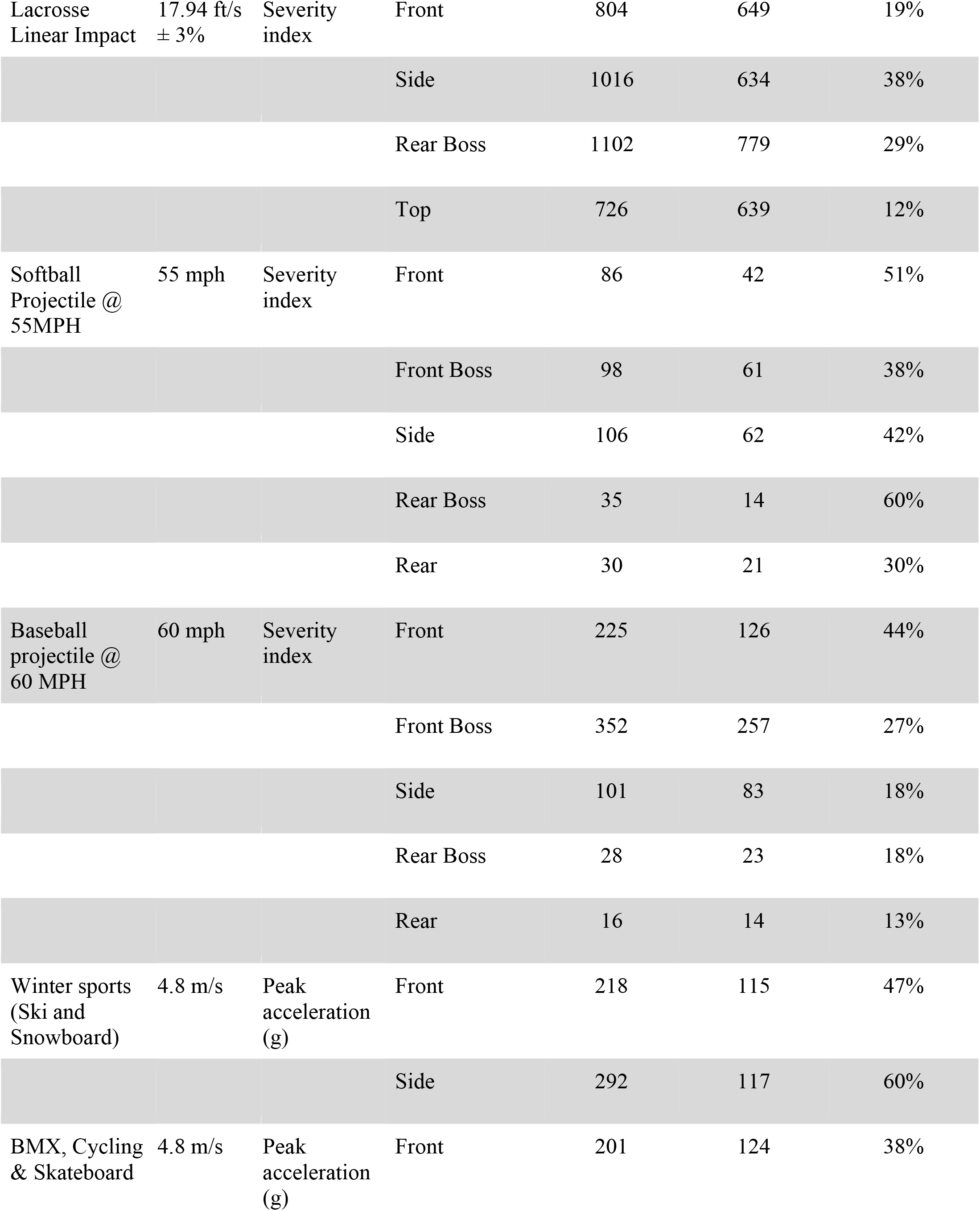
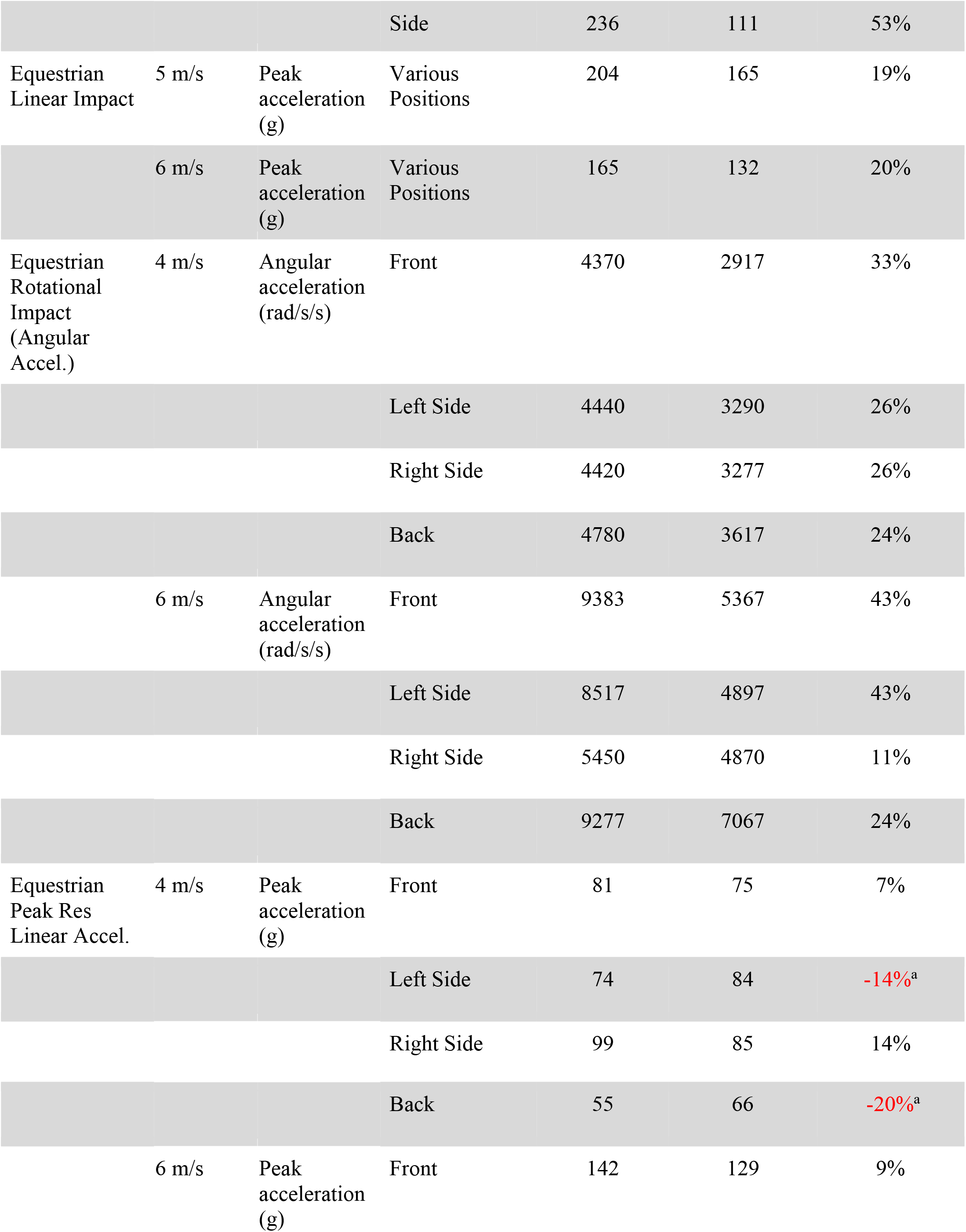
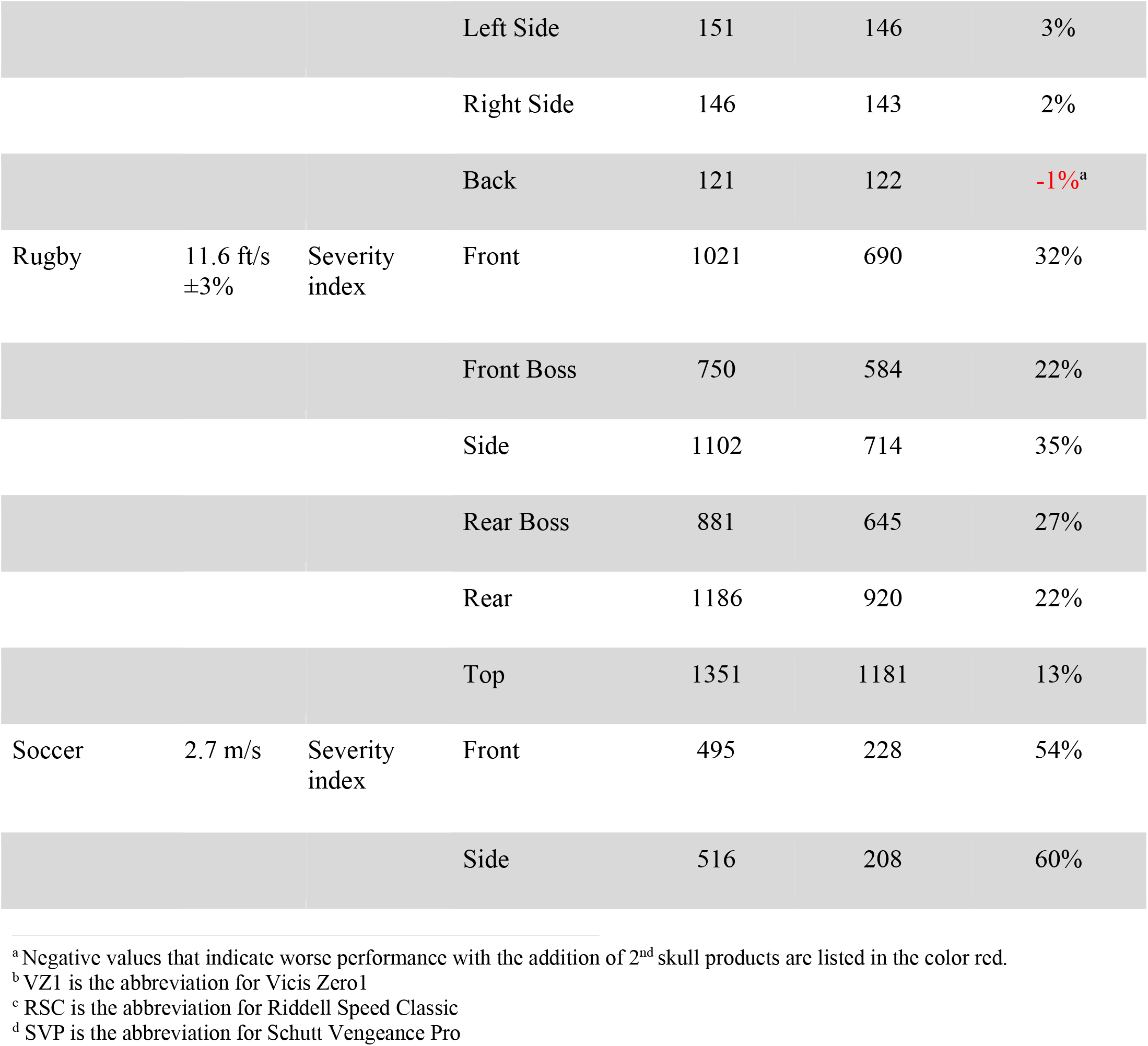
Comparison of impact severity with and without 2^nd^ Skull products.

The effectiveness of the 2^nd^ Skull Scrum Cap to reduce the severity index of head hits was tested at a single laboratory for rugby simulations. All six areas of impact (front, front boss, side, rear boss, rear and top) showed a reduction in severity index where the top of the head showed the least improvement by 13% and the highest at the side by 35% compared to hits on bare headforms.

2^nd^ Skull headbands were tested on force improvement (severity index) using soccer simulations at a single laboratory on two areas of impact (front and side) where the addition of the headband resulted in 54% and 60% improvement respectively.

2^nd^ Skull Caps were also tested for the following simulated sports:

Football simulations were conducted in two separate laboratories to measure the severity index, angular and linear acceleration and head injury criteria (HIC) in six areas of impact (front, front boss, side, rear, rear boss and top). Results showed moderate improvement in all tests (between 2% to 8%) with the addition of the 2^nd^ Skull cap.

Hockey was tested in two laboratories to measure the severity index of linear impact and average angular acceleration from rotational impact. Four areas of impact (front, side, rear and top) were tested for hockey simulations. Improvement was observed in all tests ranging from 14% at the left side of the head from rotational impact to 37% at the top of the head from linear impact.

Severity index from linear impact was tested on four areas of impact for lacrosse simulation at one laboratory. All four areas benefited from the addition of the 2^nd^ Skull Cap with improvements ranging from 12% at the top of the head to 38% at the side.

For softball and baseball simulations, the severity index from projectile impact was collected at a single laboratory on five areas of impact (front, front boss, side, rear boss and rear). Improvements were observed at all areas of impact for both softball and baseball simulations, ranging between 30% at rear and 60% at rear boss for softball and 13% at the rear and 44% at the front of the head for baseball.

For ski/snowboard and BMX/cycling/skateboard simulations, peak acceleration (g) was measured at a single testing site for two areas of impact (front and side). The 2^nd^ Skull Cap reduced impact on all tests for ski/snowboard (front=47% and side=60%) and BMX/cycling/skateboard (front=38% and side=53%).

Peak (g) was also measured for equestrian simulations in one laboratory which included linear impact test on various positions using 5 and 6m/s velocity. Peak angular acceleration (rad/s/s) and peak linear acceleration together were also measured for rotational impact tests to four areas of impact (front, left side, right side and back) using 4 and 6m/s velocity. Results on angular acceleration and most cases of linear acceleration showed improvement with the 2^nd^ Skull cap, but increased impact measures were recorded from the rotational impact test measuring linear acceleration at the left side (−14%) and back of the head (−20%) at 4m/s velocity and back of the head (−1%) at 6m/s velocity.

## Discussion

Advances in technology and simulation models have provided valuable means to assess performance characteristics of protective gears and the severity of impact sustained from sports-related impacts (Gwin et al., 2009). Using the current standard helmet impactor, four third-party laboratories tested whether supplemental 2^nd^ Skull products reduced the effects of impact to different locations of the head in comparison to the level of impact monitored with helmets alone or on bare headforms. Additional tests were completed for football given the extensive interest surrounding head injuries in the field. Although there are subtle variabilities in the level of improvement for each simulation, the results display a general pattern of reduced impact reaching the head with the addition of 2^nd^ Skull products.

Looking more specifically at each type of applied force, the majority of simulated sports showed added linear and projectile protection with the use of 2mm Skull products. Linear impact at the top of the head yielded the most improvement in football by 20% and hockey by 37% with the added 2^nd^ Skull cap. Brain injuries from linear acceleration often cause sudden changes in intracranial pressure and are thereby considered essential to mitigate. In extreme cases, athletes can experience skull fractures, which can generate a cascade of secondary injuries such as epidural hematoma, contusions and even death (Kleiven, 2013). Reducing impact severity on the top area of the head may also be of particular importance. A study investigating concussion outcomes by impact location in high school football players found significantly higher rates of loss of consciousness when the athletes sustained a concussion from impact to the top (8.0%) than other areas of the head (3.5%) (*P*=0.008) (Wilson et al., 2014). Consequences of impact to the front of the head have also been greatly discussed as the most frequent area of impact in many sports (Crisco et al., 2010; Comstock et al., 2014). For example, there are a considerable number of reports on projectile impact via a bat or ball to the front of the head in baseball and softball catchers (Beyer et al., 2012; Cusimano et al., 2017). This 2^nd^ Skull study showed reduced impact at the front of the head for both baseball by 44% and softball by 51%, which exhibits the promising effect of the 2^nd^ Skull caps in confronting this issue. These positive outcomes may be attributed to the material integrated in all 2^nd^ Skull products. Similar to standard helmets with an inner layer of energy absorption material, the supplemental 2^nd^ Skull layer may be prolonging the duration of energy reaching the head thereby dissipating the momentum caused by contact or collision (Pellman et al., 2006; Daneshvar et al., 2011). In addition to energy absorption functions produced by conventional protective gears, 2^nd^ Skull products incorporate unique lightweight XRD material made from urethane molecules that instantaneously harden with impact. The potential effectiveness of this material warrants further investigation to determine whether the results are consistent with head impact exposure on the field.

Some of the data from the equestrian simulations however did not show the same improvement pattern from linear acceleration as other sports. While the combination of 2^nd^ Skull caps and equestrian helmets may not have consistently been successful at attenuating impact from linear acceleration on the side and back of the head in the rotational impact test, favorable outcomes were recorded in all areas of impact of rotational acceleration. As the test specifically simulating linear impact showed improvement in all impacted areas, and given that a previous study evaluating the predictive value of head impact models found angular acceleration to correlate more strongly with equestrian head injury related parameters, the effectiveness of 2^nd^ Skull caps to reduce the risk of jockey head injuries should not be dismissed (Rueda et al., 2011).

In contact sports, the biomechanics of concussions have found rotational and other forces to be more damaging to the brain and thereby serve as better indicators of TBI (King et al., 2003). With increasing awareness on the detrimental effects of rotational kinematics, helmets designed for player-to-player contact sports have evolved to address parameters beyond linear acceleration. Nevertheless, in comparison to when contact sport helmets (for football and hockey) are used alone, results from this study reveal greater reduction of angular acceleration when 2^nd^ Skull caps and helmets are used together. The overall results of the three distinct helmets from the Biokinetics’s lab further acknowledge the effectiveness of 2^nd^ Skull products for various types of football helmets. 2^nd^ Skull caps offer increased protection from rotational forces for a variety of helmet types across and within sport type. We speculate that the added independent layer creates a slip pane upon rotational impact and by doing so reduces the amount of energy transferred to the head. Given the effectiveness of mitigating rotational kinematics, further research should be conducted to confirm the underlying mechanisms.

Rugby poses an additional challenge with remarkably high incidence of head injuries, as helmets are not required during play. The extent of this issue is depicted in a study directly comparing injury risk of American football and rugby. Data on concussions revealed rugby players (2.5/1000 athlete exposures) to have significantly higher rates than football players (1.4/1000 athlete exposures), which could be attributed to the different levels of protection each sport requires (Willigenburg et al., 2016). The high injury rates in rugby are also supported in a previous study, which revealed that conventional padded headgear (fabric or leather) did little to reduce the rate of concussions (Marshall et al., 2005). Given the barriers in protecting current rugby players from injury, the decline in severity index with 2^nd^ Skull scrum caps in all tested areas of impact demonstrate great potential. However, real world effects of scrum caps remain controversial as some researchers argue that added protection can change the athletes’ style of play to be more aggressive (McIntosh et al., 2003). While the simulation results are promising, further investigation is needed to confirm the efficacy of 2^nd^ Skull scrum caps during real world games.

Unlike motor vehicle accidents, the likelihood of head impact repetition is high in sports as many players return to play after recovery. Within sports, special attention has been given to severe closed-head injuries often associated with contact sports, where a single blow to the head can induce detectable impairments. Recent literatures however underscore the consequences of sub-concussive blows from sports that are less associated with high magnitude single head trauma such as soccer and lacrosse. Using TMS, Di Virgilio et al. conducted a study on female soccer players to monitor changes in brain function, more specifically cortical inhibition, following sub-concussive hits from 20 consecutive routine soccer headings (Di Virgilio et al., 2016). Results demonstrated recognizable alteration in corticomotor inhibition as well as a decline in memory test performance immediately after exposure to the headings. Another study examining the effects of sub-concussive blows to the default mode network in college rugby players found that even with acute hits below the threshold of a concussive episode, fMRI scans presented significant alterations in functional connectivity patterns (Johnson et al., 2014). The authors further exposed that the scans of those who previously experienced concussions displayed different characteristics from those without a concussion history. Even when clinical symptoms are transient or go undetected, significant neurophysiological changes occur in the brain. While many of the sub-concussive effects normalize after 24 hours, decreasing the impact of minor blows can attenuate the cumulative effects of RHI in the long term (Di Virgilio et al., 2016). These results together suggest the importance of addressing current misconceptions of sub-concussive hits and shift the focus from targeted prevention efforts to protective strategies applicable to a wider range of sports. Data from this study demonstrate how the addition of 2^nd^ Skull products can reduce impact severity for use with both multiple-hit helmets as applied in sports like hockey, football and lacrosse as well as single-impact helmets utilized in BMX, cycling and skiing/snowboarding (Hoshizaki et al., 2004).

The issue of scalability is also present in different levels of play. In professional settings, rigorous testing has led to meaningful improvements in helmets and protective gear. However, high cost top-performing products have not been scalable enough to address potential broader public health risk. In particular, young athletes including students and members of youth leagues are amongst those who do not have immediate access to top performing helmets. In 2009, it was estimated that 1,800,000 people in the United States participated in football, 1,500,000 of which were in junior high school, high school or non-federation school (Mueller & Colgate, 2011). This is of particular concern as DePadilla et al. found 15.1% of surveyed high school students reported experiencing at least one concussion in a year (CDC analyzed) (DePadilla et al., 2017). The problem may be even greater than is suggested given that RHI in the absence of overt concussive symptoms may also have residual and or cumulative impact (Stein et al., 2015). Youth athletes have a particular disadvantage as the developing and immature brain increases their susceptibility to head trauma and the subsequent neurobehavioral sequelae in comparison to adults (Guskiewicz & Valovich, 2011; Shrey et al., 2011). A study testing the fitting quality of football helmets on young athletes between the ages of 6-13, also found that only 16% of 55 helmets passed the criteria of being appropriately fit (Scifers et al., 2017). These conditions together stress the importance for vulnerable populations to have access to cost-effective supplemental protective gears, and this study offers one scalable solution to alleviate this challenge.

### Limitations

Results from our study show a consistent pattern in support of 2^nd^ Skull products, however the study is not without limitations. As with many studies using an experimental design, the degree to which the results can be translated into real impact remain inconclusive. For instance, a recent study investigating whether soccer headgear can reduce the incidence or severity of concussions demonstrated no differences between the group that wore protective headgear and the control group without any soccer headgear (McGuine et al., 2019). Purely linear or rotational acceleration independently can facilitate substantial injuries to the head including the brain. However, real world sports-related head injuries are sustained from a combination of oblique forces. The complex profiles of these forces make biomechanics replication difficult. As a commonly applied measure to assess risk of mechanical impact, HIC was reported for football. However, future studies should also include injury related parameters like HIC for other sports simulations and not exclusively focus on football. Specific to team sports, impact profiles have been suggested to change by position (Baugh et al., 2015; Barber et al., 2017). Thus, data from one model for each sport is not sufficient to draw conclusion about all positions in team sports.

Other limitations include subtle variability in methodology by testing site. While 2^nd^ Skull acted as the epicenter for these testing sites, four sites were involved in conducting the test trials. As indicated in **Table 2**, the number of trials conducted for each location and sport was different. Since the average score of these trials are reported in this study, it is possible that having increased number of trials can affect the data on level of improvement. Some of the reported data are also results of one trial which may not be sensitive enough to capture potential anomalies. Additionally, only one size 2^nd^ Skull caps, scrums and headbands were tested and did not account for different headform sizes. Overall, a larger test series would be needed to draw relevant conclusions. Given these limitations, the results of this study should be interpreted conservatively.

The products were held to rigorous testing that adhered to current gold standard assessments, but it should be noted that the authors of the paper do not have expertise in standard simulation assessments.

## Conclusion

With limited information available on the effects of RHI during life, proactive measures to minimize the severity of any head impact is recommended. While complete mitigation of sports-related brain injuries is currently limited to no play, 2^nd^ Skull products demonstrate promising mediating effects and have adaptable features for scalable use at all levels of sports participation.

## Data Availability

Raw data were generated at independent third-party laboratories commissioned by 2nd Skull. Data supporting the findings of this study are available within the article and its supplementary materials.

## Author Contributions

MR wrote the paper with significant input from HL and RA. MR performed the analysis of data provided by 2^nd^ Skull with supervision from HL.

